# Macrolevel Association of COVID-19 with Non-Communicable Disease Risk Factors in India

**DOI:** 10.1101/2020.12.21.20248684

**Authors:** Kiran Gaur, RS Khedar, Kishore Mangal, Arvind K Sharma, Rajinder K Dhamija, Rajeev Gupta

**Affiliations:** Department of Statistics, Mathematics and Computer Science, Govt SKN Agriculture College, SKN Agriculture University, Jobner, Jaipur, India; Departments of Medicine and Critical Care, Eternal Hospital, Eternal Heart Care Centre & Research Institute, Jaipur, India; Department of Community Medicine, RUHS College of Medical Sciences, Jaipur, India; Department of Neurology, Lady Hardinge Medical College, New Delhi, India; Academic Research Development Unit, Rajasthan University of Health Sciences, Jaipur, India

**Keywords:** Diabetes, Epidemiology, Hypertension, Non-communicable diseases, SARS CoV-2, Social determinants, Vaccination

## Abstract

**Objective:** Greater COVID-19 related mortality has been reported among persons with various non-communicable diseases (NCDs). We performed an ecological study to determine the association of state-level cases and deaths with NCD risk factors and healthcare and social indices.

**Methods:** We obtained cumulative national and state-level data on COVID-19 cases and deaths from publicly available database www.covid19india.org from February to end November 2020. To identify association with major NCD risk factors, NCDs, healthcare related and social variables we obtained data from public sources. Association was determined using univariate and multivariate statistics.

**Results:** More than 9.5 million COVID-19 cases and 135,000 deaths have been reported in India at end November 2020. There is significant positive correlation (Pearson’s r) of state-level COVID-19 cases and deaths per million, respectively, with NCD risk factors- obesity (0.64, 0.52), hypertension (0.28, 0.16), diabetes (0.66, 0.46), literacy, NCD epidemiological transition index (0.58, 0.54) and ischemic heart disease mortality (0.22, 0.33). Correlation is also observed with indices of healthcare access and quality (0.71, 0.61), urbanization (0.75, 0.73) and human (0.61, 0.56) and sociodemographic (0.70, 0.69) development. Multivariate adjusted analyses shows strong correlation of COVID-19 burden and deaths with NCD risk factors (r^2^=0.51, 0.43), NCDs (r^2^=0.32, 0.16) and healthcare related factors (r^2^=0.52, 0.38).

**Conclusions:** COVID-19 disease burden and mortality in India is ecologically associated with greater state-level burden of NCDs and risk factors, especially obesity and diabetes.

**KEY MESSAGES:**

- There is significant state-level variability in COVID-19 cases and deaths in India.
- In a macrolevel statistical analysis we find that Indian states with better human and sociodemographic indices, more literacy, longer age, greater burden of non-communicable diseases and risk factors have greater COVID-19 case burden and mortality.
- Non-communicable disease risk factors- obesity and diabetes are the most important determinants on multivariate analyses.

## INTRODUCTION

There is strong association of COVID-19 with non-communicable diseases (NCDs) and their risk factors.^1^ Multiple studies from across the globe have reported that individuals with established coronary heart disease, heart failure, chronic respiratory, renal or liver disease and cancers or their risk factors such as diabetes, hypertension, obesity, and other vascular risk factors are at greater risk of acquiring infection and developing complications and deaths from COVID-19.^2-10^ Environmental factors such as urbanization, crowding, ambient and indoor air pollution and poor sanitation and low socioeconomic status are also important in increasing the risk of disease and deaths.^5,11-13^

India has one of the highest absolute burden of COVID-19 infection with more than 9.5 million cases and 135,000 deaths reported by November 2020.^14^ However, the rates of infection related cases and deaths per million population are lower than many developed countries.^15^ Mortality rate (case-fatality rates) from the infection are also lower than many developed countries.^15^ There is also significant state-level variation in incidence of cases and deaths with more developed and urbanized states of the country reporting greater disease burden.^16,17^ Small case-series from India have reported greater mortality from COVID-19 among hospitalized patients with diabetes, heart failure, chronic respiratory and chronic liver and kidney diseases.^18-20^ It is likely that state-level variation in incidence of COVID-19 and mortality is influenced by NCD and their risk factors as hypothesized in a previous study.^21^ We performed a macrolevel ecological analysis using publicly available data to identify association of regional (state-level) burden of COVID-19 cases and deaths with non-communicable diseases, NCD risk factors and selected social determinants.

## METHODS

The study has been conducted using publicly available data on COVID-19, NCD risk factors and social factors.^14,22^ The project proposal was approved by the institutional ethics committee as part of COVID-19 database at our centre. Daily data on COVID-19 in various states and regions of India are being regularly updated at a non-commercial public website, www.covid19india.org.^14^ This website updates daily data on COVID-19 related cases, deaths, recovery and testing at the state level of India. We obtained data for all the states in the country and clubbed daily data into weekly and monthly numbers beginning February 2020 to end of November 2020. The data were collated on spreadsheets. We then calculated number of cases and deaths per million population, using 2020 estimates, for each state (Table 1).

**Table 1:** COVID-19 cases, deaths and rates/million population in various states in India at 30^th^November 2020.

| States | Population<br>2020 | Cases |  | Deaths |  | Case:Fatality<br>ratio |
| --- | --- | --- | --- | --- | --- | --- |
|  |  | Number | Per million | Number | Per million |  |
| India | 1371360650 | 9463254 | 6900.63 | 137659 | 100.38 | 1.45 |
| Andhra Pradesh | 53903393 | 868064 | 16104.07 | 6992 | 129.71 | 0.81 |
| Arunachal Pradesh | 1570458 | 16282 | 10367.68 | 54 | 34.38 | 0.33 |
| Assam | 35607039 | 212776 | 5975.67 | 981 | 27.55 | 0.46 |
| Bihar | 124799926 | 235616 | 1887.95 | 1264 | 10.13 | 0.54 |
| Chhattisgarh | 29436231 | 237322 | 8062.24 | 2861 | 97.19 | 1.21 |
| Delhi | 18710922 | 570374 | 30483.48 | 9174 | 490.30 | 1.61 |
| Goa | 1586250 | 47963 | 30236.72 | 688 | 433.73 | 1.43 |
| Gujarat | 63872399 | 209780 | 3284.36 | 3989 | 62.45 | 1.90 |
| Haryana | 28204692 | 234126 | 8300.96 | 2428 | 86.08 | 1.04 |
| Himachal Pradesh | 7451955 | 40518 | 5437.23 | 635 | 85.21 | 1.57 |
| Jammu Kashmir | 13606320 | 110224 | 8100.94 | 1694 | 124.50 | 1.54 |
| Jharkhand | 38593948 | 109151 | 2828.19 | 964 | 24.98 | 0.88 |
| Karnataka | 67562686 | 884897 | 13097.42 | 11778 | 174.33 | 1.33 |
| Kerala | 35699443 | 602983 | 16890.54 | 2245 | 62.89 | 0.37 |
| Madhya Pradesh | 85358965 | 206128 | 2414.84 | 3260 | 38.19 | 1.58 |
| Maharashtra | 123144223 | 1823896 | 14811.06 | 47151 | 382.89 | 2.59 |
| Manipur | 3091545 | 25045 | 8101.13 | 281 | 90.89 | 1.12 |
| Meghalaya | 3366710 | 11810 | 3507.88 | 111 | 32.97 | 0.94 |
| Mizoram | 1239244 | 3825 | 3086.56 | 5 | 4.03 | 0.13 |
| Nagaland | 2249695 | 11186 | 4972.23 | 64 | 28.45 | 0.57 |
| Odisha | 46356334 | 318725 | 6875.54 | 1792 | 38.66 | 0.56 |
| Punjab | 30141373 | 152091 | 5045.92 | 4807 | 159.48 | 3.16 |
| Rajasthan | 81032689 | 268063 | 3308.08 | 2312 | 28.53 | 0.86 |
| Sikkim | 690251 | 4990 | 7229.25 | 109 | 157.91 | 2.18 |
| Tamilnadu | 77841267 | 781915 | 10044.99 | 11712 | 150.46 | 1.50 |
| Telangana | 39362732 | 269816 | 6854.61 | 1458 | 37.04 | 0.54 |
| Tripura | 4169794 | 32692 | 7840.20 | 367 | 88.01 | 1.12 |
| Uttar Pradesh | 237882725 | 543888 | 2286.37 | 7761 | 32.63 | 1.43 |
| Uttarakhand | 11250858 | 74795 | 6647.94 | 1231 | 109.41 | 1.65 |
| West Bengal | 99609303 | 483484 | 4853.80 | 8424 | 84.57 | 1.74 |

We obtained data on state-level burden of related NCD risk factors (obesity, hypertension, diabetes, smoking, literacy), NCDs (epidemiological transition index, ETI, proportion of disability adjusted life years due to NCDs vs. communicable, maternal, neonatal and nutritional diseases), and diseases (ischemic heart disease deaths and disability adjusted life years, DALYs) from national data sources as shown in Table 2.^23-29^ Data on healthcare related factors (healthcare availability index (HAI), healthcare access and quality in index (HAQI) and sociodemographic indices of each state were also obtained from public websites.^25,26^ The following indices were used: urbanization index (UI, proportion of urban to rural population),^27^ human development index (HDI),^28^ sociodemographic index (SDI),^25^ social development index,^29^ and vulnerability index.^30^ Details of estimation of each of these indices have been reported earlier.^22^

**Table 2:** State level non-communicable disease risk factors and burden, healthcare indices and social factors.

|  | Smoking/<br>Tobacco (%) | Obesity (%) | Hypertension (%) | Diabetes (%) | Literacy (%) | Epidemiological<br>Transition index | IHD DALYs | IHD death<br>rate/100,000 | Healthcare<br>availability index | Healthcare access<br>and quality index | Urbanization index | Human development<br>index | Socio Demographic<br>Index | Social Development<br>Index | Vulnerability Index |
| --- | --- | --- | --- | --- | --- | --- | --- | --- | --- | --- | --- | --- | --- | --- | --- |
| Data sources | NFHS | NFHS | DLHS | DLHS | Census | GBD | GBD | GBD | NITI | GBD | Census | Niti | GBD | IEG | IIPS |
| Andhra Pradesh | 26.8 | 33.1 | 24.3 | 9.1 | 67.02 | 2.70 | 2099143 | 163.43 | 65.13 | 46.50 | 33.49 | 0.64 | 0.59 | 0.652 | 0.71 |
| Arunachal Pradesh | 60.0 | 18.5 | 23.0 | 4.1 | 65.39 | 1.82 | 14399 | 37.27 | 46.07 | 44.30 | 22.67 | 0.66 | 0.59 |  | 0.03 |
| Assam | 63.9 | 12.5 | 18.2 | 3.5 | 72.19 | 1.61 | 573640 | 66.41 | 48.85 | 34.00 | 14.08 | 0.61 | 0.56 | 0.632 | 0.26 |
| Bihar | 50.1 | 11.2 | 18.9 | 2.5 | 61.8 | 1.35 | 2666743 | 103.02 | 32.11 | 37.00 | 11.30 | 0.58 | 0.44 | 0.226 | 0.97 |
| Chhattisgarh | 55.2 | 11.0 | 15.2 | 4.4 | 70.28 | 1.67 | 649384 | 93.15 | 53.36 | 37.40 | 23.24 | 0.61 | 0.56 | 0.543 | 0.31 |
| Delhi | 30.4 | 25.8 | 21.9 | 8.2 | 86.21 | 2.63 | 516942 | 107.62 | 49.42 | 56.20 | 97.50 | 0.75 | 0.75 |  | 0.51 |
| Goa | 20.8 | 32.2 | 28.8 | 16.7 | 88.7 | 4.76 | 45319 | 136.28 | 51.90 | 64.80 | 62.17 | 0.76 | 0.74 |  | 0.31 |
| Gujarat | 51.4 | 19.9 | 0.0 | 0.0 | 78.03 | 2.17 | 2508489 | 159.59 | 63.52 | 45.00 | 42.58 | 0.67 | 0.62 | 0.635 | 0.77 |
| Haryana | 35.8 | 18.0 | 21.3 | 4.9 | 75.55 | 2.50 | 1213832 | 174.92 | 53.51 | 45.00 | 34.79 | 0.71 | 0.66 |  | 0.40 |
| Himachal Pradesh | 40.5 | 24.6 | 33.7 | 3.2 | 82.8 | 3.33 | 179914 | 113.7 | 62.41 | 51.70 | 10.04 | 0.72 | 0.67 | 0.499 | 0.06 |
| Jammu Kashmir | 38.2 | 22.1 | 0.0 | 0.0 | 67.16 | 2.94 | 401570 | 143.21 | 62.37 | 46.70 | 27.21 | 0.69 | 0.60 | 0.640 | 0.71 |
| Jharkhand | 48.6 | 10.1 | 20.6 | 3.3 | 66.41 | 1.45 | 790885 | 95.69 | 51.33 | 37.40 | 24.05 | 0.60 | 0.52 | 0.921 | 0.91 |
| Karnataka | 34.3 | 22.1 | 22.6 | 9.5 | 75.37 | 2.94 | 2577991 | 169.35 | 61.14 | 46.60 | 38.57 | 0.68 | 0.61 | 0.468 | 0.57 |
| Kerala | 25.7 | 33.0 | 40.3 | 14.2 | 94 | 6.25 | 1227249 | 170.36 | 74.01 | 63.90 | 47.72 | 0.78 | 0.68 | 0.730 | 0.31 |
| Madhya Pradesh | 59.5 | 11.4 | 17.0 | 2.4 | 69.32 | 1.67 | 2283586 | 122.41 | 38.39 | 39.50 | 27.63 | 0.61 | 0.53 |  | 1.00 |
| Maharashtra | 36.5 | 21.9 | 23.2 | 5.0 | 82.34 | 3.03 | 4469799 | 163.78 | 63.99 | 49.80 | 45.23 | 0.70 | 0.67 |  | 0.83 |
| Manipur | 70.6 | 22.6 | 21.3 | 7.6 | 76.9 | 2.38 | 50620 | 68.98 | 60.60 | 44.20 | 30.21 | 0.69 | 0.61 |  | 0.54 |
| Meghalaya | 72.2 | 11.0 | 20.4 | 2.9 | 74.43 | 1.56 | 29460 | 39.41 | 55.95 | 39.60 | 20.08 | 0.66 | 0.59 |  | 0.29 |
| Mizoram | 80.4 | 20.5 | 19.2 | 3.4 | 91.33 | 1.89 | 8193 | 27.37 | 74.97 | 48.90 | 51.51 | 0.71 | 0.63 |  | 0.14 |
| Nagaland | 69.4 | 14.1 | 33.2 | 5.6 | 79.6 | 2.13 | 28504 | 50.92 | 38.51 | 46.10 | 28.97 | 0.68 | 0.66 | 0.467 | 0.66 |
| Odisha | 55.9 | 16.5 | 16.1 | 3.2 | 72.89 | 1.72 | 808170 | 71.55 | 35.97 | 36.30 | 16.68 | 0.61 | 0.55 | 0.766 | 0.80 |
| Punjab | 19.2 | 26.5 | 31.7 | 6.3 | 75.84 | 3.45 | 1725787 | 261.05 | 63.01 | 49.50 | 37.49 | 0.72 | 0.65 |  | 0.43 |
| Rajasthan | 46.9 | 12.7 | 18.6 | 2.7 | 66.11 | 1.52 | 1762007 | 94.96 | 43.10 | 40.70 | 24.89 | 0.63 | 0.53 | 0.732 | 0.69 |
| Sikkim | 40.3 | 27.4 | 29.7 | 4.5 | 81.42 | 2.22 | 10289 | 65.07 | 50.51 | 50.50 | 24.97 | 0.72 | 0.63 | 0.732 | 0.00 |
| Tamilnadu | 31.7 | 29.4 | 23.2 | 15.3 | 80.09 | 3.85 | 3629123 | 207.5 | 60.41 | 51.20 | 48.45 | 0.71 | 0.65 |  | 0.57 |
| Telangana | 28.3 | 26.0 | 22.9 | 8.0 |  | 2.63 | 1213030 | 134.51 | 59.00 | 48.50 | 48.45 | 0.67 | 0.61 | 0.340 | 0.94 |
| Tripura | 67.8 | 15.3 | 19.8 | 9.1 | 87.22 | 2.22 | 112459 | 106.59 | 46.38 | 42.30 | 26.18 | 0.66 | 0.57 |  | 0.63 |
| Uttar Pradesh | 53.0 | 13.2 | 17.7 | 3.2 | 67.68 | 1.47 | 5008099 | 99.47 | 28.61 | 34.90 | 22.28 | 0.60 | 0.51 | 0.709 | 0.89 |
| Uttarakhand | 43.7 | 18.4 | 26.3 | 3.8 | 78.82 | 2.17 | 297063 | 119.58 | 40.20 | 43.20 | 30.55 | 0.68 | 0.64 |  | 0.43 |
| West Bengal | 58.8 | 16.2 | 20.7 | 9.2 | 76.26 | 3.03 | 3284771 | 146.32 | 57.17 | 47.10 | 31.87 | 0.64 | 0.58 |  | 0.83 |
| DLHS District level health survey; GBD Global burden of diseases study; IEG Institute for economic growth; IIPS Indian institute of population sciences; NFHS National family health survey-4; NITI National institute for transforming India. |  |  |  |  |  |  |  |  |  |  |  |  |  |  |  |

### Statistical analyses

We determined burden of COVID-19 cases and deaths per million population at end of November 2020 for each state of the country. Descriptive statistics for each state is reported. To determine association of COVID-19 cases and death with state-level NCD and risk factors, healthcare indices and sociodemographic variables we calculated Pearson’s correlation coefficient (r value) using SPSS Statistical Package. MS Office Powerpoint-2007 was used to plot scatter-graphs for estimation of correlation of various NCD risk factors, NCD and sociodemographic indices with total cases/million and deaths/million. Linear correlation was calculated and R^2^ values are reported.). To identify factors of importance we also calculated multivariate regression statistics. P value <0.5 was considered significant.

## RESULTS

More than 9.5 million COVID-19 cases and 135,000 deaths have been reported in India at end of November 2020.^14^ The national burden of cases and deaths is 6900/million and 100.4/million respectively. There are wide disparities in rates of cases and deaths in different states of India (Table 1) with reported cases more than 20,000/million in states of Delhi and Goa and 10,000-20,000/million in a number of states. Similarly deaths rates of more than 300/million are observed in Delhi (490), Goa (434) and Maharashtra (383). The case-fatality rate also shows significant differences with less than 0.5% in Mizoram, Arunachal Pradesh, Kerala and Assam to more than 2% in Punjab, Maharashtra and Sikkim. The data on various state-level NCD risk factors (obesity, hypertension, diabetes, smoking, and literacy), NCDs (NCD epidemiological transition index, ischemic heart disease), healthcare indices and various sociodemographic indices are in Table 2 and shows wide variability.

Univariate correlation analysis (Pearson’s r value) (Table 3) shows significant correlation of state-level COVID-19 cases/million with obesity (0.642), hypertension (0.283), diabetes (0.656) and literacy (0.460) and inverse association with tobacco use (−0.555). Significant correlation is also observed with ETI (0.585), HAQI (0.710), urbanization (0.745), HDI (0.608), sociodemographic index (0.698) and social development index (0.608). COVID-19 death rates have significant correlation with obesity (0.520), diabetes (0.458) and literacy (0.458) and inverse association with tobacco use (−0.554). Significant correlation is also observed with ETI (0.437), IHD death rate (0.332), HAQI (0.614), urbanization (0.726), HDI (0.563) and sociodemographic index (0.686). There is significant association of state-level cases/million as well as deaths/million (r=0.86). Correlation of NCD risk factors (obesity, hypertension, diabetes and tobacco with COVID-19 cases (Figure 1) and deaths (Figure 2) show that the strongest association is with state-level diabetes prevalence.

**Table 3:** Association of various non-communicable diseases, healthcare and social indices with COVID-19 cases and deaths (Pearson’s r)

| Variables | Cases/million | Deaths/million |
| --- | --- | --- |
| **Non-communicable disease risk factors** |  |  |
| Obesity (NFHS) | 0.642** | 0.520** |
| Hypertension (DLHS/AHS) | 0.283 | 0.164 |
| Diabetes (DLHS/AHS) | 0.656** | 0.458* |
| Literacy rates (Census) | 0.460* | 0.458* |
| Smoking /Tobacco (NFHS) | -0.555** | -0.554** |
| **Non-communicable diseases** |  |  |
| NCD Epidemiological transition index (GBD) | 0.585** | 0.437* |
| Ischemic heart disease DALYs (GBD) | -0.120 | 0.067 |
| Ischemic heart disease death rate (GBD) | 0.224 | 0.332* |
| **Healthcare Related Factors** |  |  |
| Healthcare availability index (NITI) | 0.226 | 0.177 |
| Healthcare accessibility and quality index (GBD) | 0.710** | 0.614** |
| **Macrolevel Social Factors** |  |  |
| Urbanization index (Census) | 0.745** | 0.726** |
| Human development index (MOSPI) | 0.608** | 0.563** |
| Sociodemographic index (GBD) | 0.698** | 0.686** |
| Vulnerability index (IIPH) | -0.207 | -0.095 |
| Social development index (IEG) | 0.608* | 0.035 |
| *p<0.05, **p<0.01, ***p<0.001<br>DLHS/AHS District level health survey/Annual health survey; GBD Global burden of diseases study; IEG Institute for Economic Growth; IIPH Indian institute of population health; MOSPI Ministry of Statistics and Program Implementation; NCD non-communicable diseases; NFHS National family health survey-4; NITI National institute for transforming India; |  |  |

**Figure 1:**
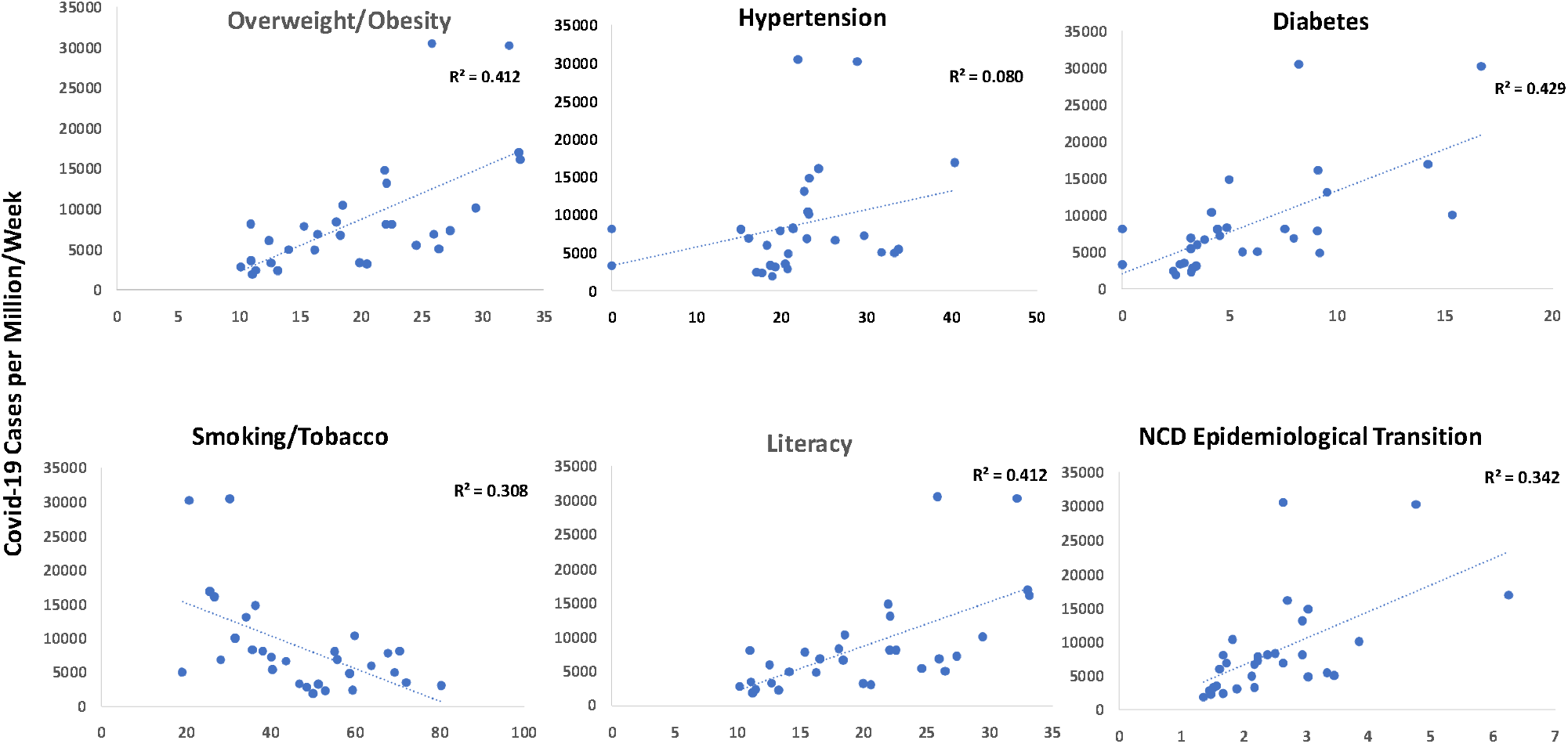
Association of multiple non-communicable disease (NCD) risk factors with cumulative COVID-19 cases/million.

**Figure 2:**
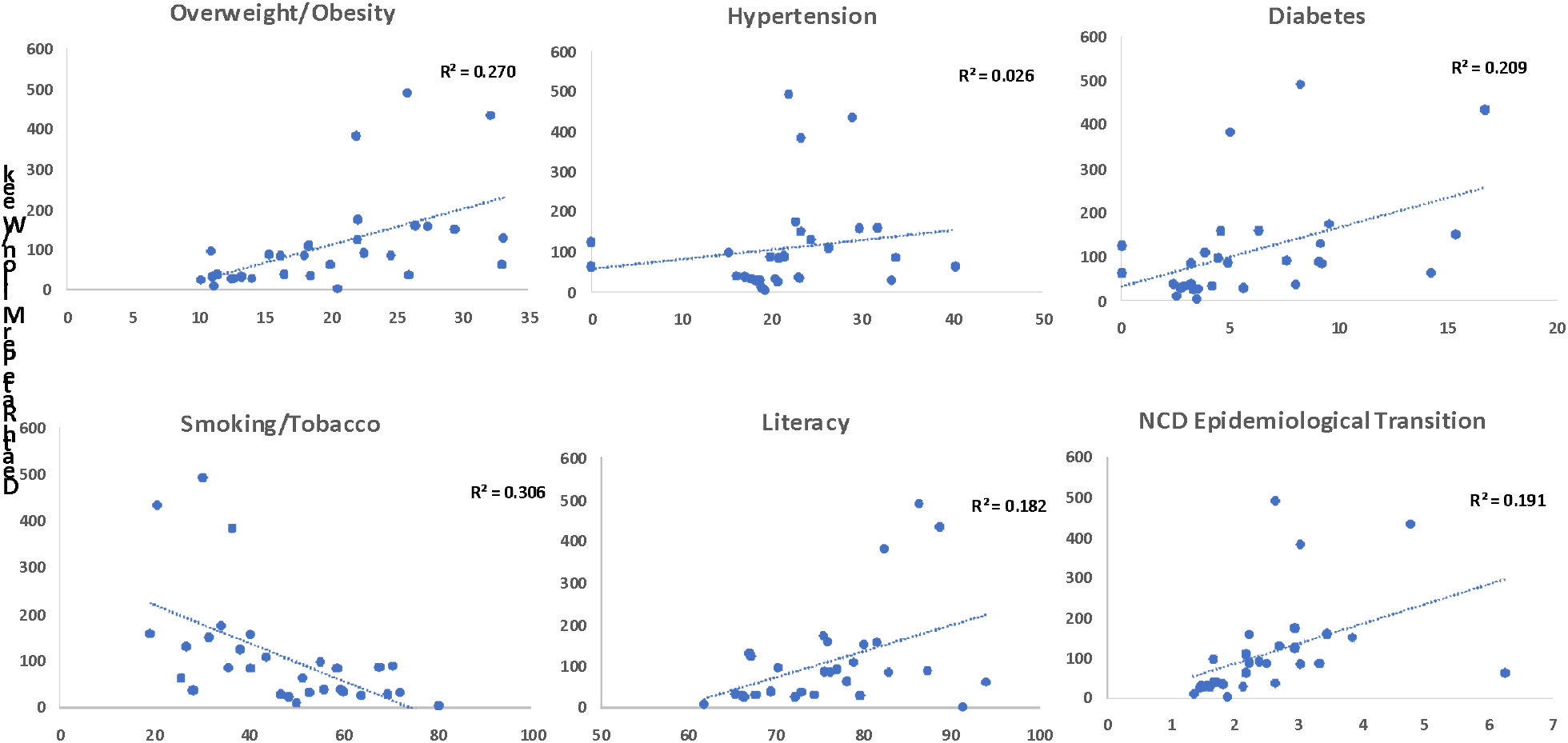
Association of non-communicable disease (NCD) risk factors with cumulative COVID-19 deaths/million.

Multivariate analyses shows significant association of COVID-19 cases as well as deaths with NCDs risk factors (unadjusted r^2^, cases 0.73, deaths 0.51; multivariate adjusted r^2^, cases 0.65, deaths 0.36) (Table 4). NCD burden as estimated by epidemiological transition index shows significant association with COVID-19 cases (unadjusted r^2^ 0.34, adjusted r^2^ 0.32) and not with deaths. This association disappears after adjustment for NCD risk factors. Association of COVID-19 cases and deaths was also observed with healthcare related factors (unadjusted r^2^, cases 0.55, deaths 0.42; multivariate adjusted r^2^, cases 0.52, deaths 0.38), however, this association was no longer observed with various social variables after accounting for the NCD risk factors, NCDs and healthcare related factors.

**Table 4:** Multiple regression analysis of COVID-19 cases and deaths with various risk factors.

| Variables | COVID-19 Cases/million |  |  | COVID-19 Deaths/million |  |  |
| --- | --- | --- | --- | --- | --- | --- |
|  | B value | Standardized beta | P value | B value | Standardized beta | P value |
| **Non-communicable disease risk factors** |  |  |  |  |  |  |
| Overweight or Obesity | 109.90 | 0.11 | 0.66 | -2.29 | -0.13 | 0.62 |
| Hypertension | -172.84 | -0.20 | 0.23 | -3.94 | -0.27 | 0.14 |
| Diabetes | 747.18 | 0.43 | 0.04 | 5.51 | 0.19 | 0.37 |
| Smoking/ Tobacco | -150.87 | -0.34 | 0.11 | -4.66 | -0.63 | 0.01 |
| Literacy rate | 191.32 | 0.22 | 0.25 | 6.48 | 0.44 | 0.04 |
| Multivariate correlation | $r^2=0.601$; Adjusted $r^2=0.514$ | | | $r^2=0.533$, Adjusted $r^2=0.433$ | | |
| **Non-communicable diseases burden** |  |  |  |  |  |  |
| NCD epidemiological transition index | 3907.961 | 0.58 | <0.01 | 49.56 | 0.44 | 0.02 |
| Univariate correlation | $r^2=0.343$; Adjusted $r^2=0.319$ | | | $r^2=0.192$; Adjusted $r^2=0.163$ | | |
| Multivariate correlation | Model not significant |  |  | Model not significant |  |  |
| **Healthcare related factors** |  |  |  |  |  |  |
| Healthcare availability index | -157.77 | -0.26 | 0.111 | -2.60 | -0.25 | 0.169 |
| Healthcare accessibility and quality index | 814.88 | 0.86 | <0.001 | 12.21 | 0.76 | <0.001 |
| Multivariate correlation | $r^2=0.549$; Adjusted $r^2=0.516$ | | | $r^2=0.420$; Adjusted $r^2=0.377$ | | |
| **Macrolevel social factors** | Model Not Significant |  |  | Model Not Significant |  |  |

## DISCUSSION

Indian states with greater burden of non-communicable disease risk factors (obesity, diabetes) and in advanced stage of epidemiological transition with greater NCD burden, better healthcare access and quality, urbanization and human and sociodemographic development have greater COVID-19 case burden and mortality. This finding has important implications for implementation of population and individual level preventive measures and equitable vaccine deployment.

That NCD risk factors as well as the disease conditions increase risk of adverse outcomes in COVID-19 is well reported. Large registries as well as population based databases in UK, USA, Europe and elsewhere have reported such association. OpenSafely and QRESEARCH patient-level cohorts in UK,^5,31^ and multiple cohorts in USA have reported this association. Little is, however, known regarding macrolevel association of NCD risk factors and diseases on COVID-19 burden and mortality. A significant association of aging with adverse outcomes in COVID-19 has been reported in China, Europe and USA.^1^ In India a study reported heightened adverse events among the elderly using a modeling algorithm.^32^ It is well known that NCD risk factors such as hypertension and diabetes as well as NCDs increase with age, also in India,^33^ and this could be a reason of greater COVID-19 related deaths in more urbanized and better developed states as observed in the present study (Table 3). Average life expectancy of all these states is significantly greater than those states where the disease burden is lower.^26^ A macrolevel analysis from India previously reported greater case-fatality in states with greater aging population.^21^ In USA, a study reported macrolevel association of COVID-19 burden and case-fatality with air pollution, gross domestic product (GDP) per capita and urbanization.^34^ Another country-level macroecological study reported association of COVID-19 burden and mortality with national GDP, urbanization, population density, number of tourists in a country and geographic longitude.^35^ Influence of demographic and socioeconomic factors in the COVID-19 case-fatality rate globally was examined among 10.5 million cases in 209 countries. Doubling in size of population, proportion of female smokers, higher stringency index and active testing policies were associated with higher case-fatality rate while inverse association was found between cardiovascular disease death rate and diabetes prevalence.^36^ In the present study we did not find any association of case-fatality rates in different Indian states with various NCD risk factors and social indices (data not shown) and therefore we have not commented on this issue. No sub-national studies, similar to the present study, have been reported from developed countries although recent surge of COVID-19 in rural locations in USA, where the NCDs as well as risk factors are significantly greater,^37^ suggest such an association.^38^ More studies in different geographic locations are needed.

In the present study, association of COVID-19 with sociodemographic factors such as urbanization and other indices of development and better healthcare access and quality indices indicate better diagnosis of the condition in these locations.^1^ In lesser developed states of the country as well as rural areas of India it is likely that the condition is not diagnosed well due to lack of access to quality healthcare.^39^ Undiagnosed COVID-19 related deaths in the rural hinterlands of the country are also possible.^40^ Possible gaps in COVID-19 data from India has been highlighted.^41^ Other limitations of our analyses are absence of detailed readily available cause-specific morbidity and mortality data, lack of population based registries of COVID-19 cases and deaths, absence of concurrent mortality data to evaluate excess deaths from COVID-19 and absence of other collinear data.

Our study shows that COVID-19 burden is significantly greater in more developed and urbanized states of the country where there is a greater prevalence of NCD risk factors. These findings have important implications for prioritization of deployment of preventive strategies and vaccinations. Large scale government-led non-pharmaceutical prevention strategies that have proven effective in COVID-19 control include mandatory mask-wearing in public spaces, limited public gatherings (<100 or <10 people depending on context), closure of non-essential businesses, schools and universities, and stay at home orders with appropriate exemptions.^42^ Individual level non-pharmacological interventions include proper masking, physical distancing and avoiding crowded spaces. Although, vaccines have become available and need scientifically appropriate strategies to achieve maximum impact and development of population-level immunity.^43,44^ Equitable vaccine distribution is essential.^45^ However, in the present macrolevel scenario highlighted in this report we posit that more developed states, who are at forefront of the COVID-19 epidemic, need non-pharmacological interventions as well as vaccines more than the lesser developed.

## Data Availability

Al the data are provided in the manuscript. Tables 1 and 2 contain all data used for the present study.

## Notes

### Competing Interest Statement

The authors have declared no competing interest.

### Clinical Trial

NA

### Funding Statement

Nil.

### Author Declarations

Institutional Ethics Committee, Eternal Heart Care Centre & Research Institute, Jaipur, India.

